# A tertiary center experience of multiple myeloma patients with COVID-19: lessons learned and the path forward

**DOI:** 10.1101/2020.06.04.20122846

**Authors:** Bo Wang, Oliver Van Oekelen, Tarek H. Mouhieddine, Diane Marie Del Valle, Joshua Richter, Hearn Jay Cho, Shambavi Richard, Ajai Chari, Sacha Gnjatic, Miriam Merad, Sundar Jagannath, Samir Parekh, Deepu Madduri

## Abstract

**Background:** The COVID-19 pandemic, caused by SARS-CoV-2 virus, has resulted in over 100,000 deaths in the United States. Our institution has treated over 2,000 COVID-19 patients during the pandemic in New York City. The pandemic directly impacted cancer patients and the organization of cancer care. Mount Sinai Hospital has a large and diverse multiple myeloma (MM) population. Herein, we report the characteristics of COVID-19 infection and serological response in MM patients in a large tertiary care institution in New York.

**Methods:** We performed a retrospective study on a cohort of 58 patients with a plasma-cell disorder (54 MM, 4 smoldering MM) who developed COVID-19 between March 1, 2020 and April 30, 2020. We report epidemiological, clinical and laboratory characteristics including persistence of viral detection by polymerase chain reaction (PCR) and anti-SARS-CoV-2 antibody testing, treatments initiated, and outcomes.

**Results:** Of the 58 patients diagnosed with COVID-19, 36 were hospitalized and 22 were managed at home. The median age was 67 years; 52% of patients were male and 63% were non-white. Hypertension (64%), hyperlipidemia (62%), obesity (37%), diabetes mellitus (28%), chronic kidney disease (24%) and lung disease (21%) were the most common comorbidities. In the total cohort, 14 patients (24%) died. Older age (>70 years), male sex, cardiovascular risk, and patients not in complete remission (CR) or stringent CR were significantly (p<0.05) associated with hospitalization. Among hospitalized patients, laboratory findings demonstrated elevation of traditional inflammatory markers (CRP, ferritin, D-dimer) and a significant (p<0.05) association between elevated inflammatory markers, severe hypogammaglobulinemia, non-white race, and mortality. Ninety-six percent (22/23) of patients developed antibodies to SARS-CoV-2 at a median of 32 days after initial diagnosis. Median time to PCR negativity was 43 (range 19-68) days from initial positive PCR.

**Conclusions:** Drug exposure and MM disease status at the time of contracting COVID-19 had no bearing on mortality. Mounting a severe inflammatory response to SARS-CoV-2 and severe hypogammaglobulinemia were associated with higher mortality. The majority of patients mounted an antibody response to SARS-CoV-2. These findings pave a path to identification of vulnerable MM patients who need early intervention to improve outcome in future outbreaks of COVID-19.

## Introduction

The coronavirus disease 2019 (COVID-19) pandemic, caused by Severe Acute Respiratory Syndrome Coronavirus-2 (SARS-CoV-2), represents a world-wide public health crisis. Patient care has been drastically altered, primarily in epidemic, urban areas. As of May 22, 2020, New York City had nearly 200,000 confirmed cases of COVID-19 with over 16,000 deaths and a patient death rate of 21%^1^, with cancer patients comprising about 8% of all COVID-19 fatalities in the state of New York^2^. Mount Sinai Hospital, a tertiary care center in New York City, has treated over 2,000 admitted COVID-19 patients thus far. At our cancer center, we actively care for a large and particularly diverse population of over 3000 multiple myeloma (MM) patients. Like many other centers in the region and the world, clinical care at our institution has seen significant changes in an attempt to mitigate the spread of SARS-CoV-2 to vulnerable cancer patients receiving treatment. Balancing the competing risks of treatment delay or alteration versus infection is essential and depends upon understanding the clinical profile of COVID-19 in this vulnerable population.

Limited studies describing the impact of COVID-19 both in the United States^3^ and abroad^4-7^ suggest a higher risk of hospitalization and poor outcomes including death in certain subsets of cancer patients. The effect of COVID-19 on patients with MM, the second most common hematological malignancy, is of particularly great concern due to immunosuppression associated with the disease, and at this time remains incompletely understood. MM is a plasma cell malignancy, diagnosed at a median age around 70 years in patients often with multiple comorbidities^8^. MM is associated with both cellular and humoral immune dysfunction and causes a state of generalized immune suppression, leaving patients especially vulnerable to infections^9,10^.

In contrast to the reported immunosuppressive nature of MM, COVID-19 infection has demonstrated propensity for triggering an uncontrolled immune inflammatory cascade^11-13^ that bears resemblance to cytokine release syndrome (CRS) seen in patients treated with chimeric antigen receptor (CAR) T cells and bispecific antibodies^14,15^. Inflammatory markers and cytokines, including C-reactive protein (CRP), ferritin, interleukin (IL)-6, have been significantly elevated in multiple cohorts of patients infected with COVID-19^16-19^.

We aimed to characterize the population of MM patients at our institution who developed COVID-19 in the epicenter of the pandemic in the United States. To address this, we retrospectively analyzed a cohort of 58 MM and smoldering MM (SMM) patients treated at the Mount Sinai Hospital who were diagnosed with COVID-19 between March 1 and April 30, 2020. We have identified several demographic characteristics and comorbidities associated with hospitalization and elevation of certain inflammatory markers associated with increased mortality as described below.

## Methods

### Study design, inclusion criteria and data collection

The study was designed from a register of patients with SMM and MM in any phase of response, currently receiving treatment or follow-up at the Mount Sinai Hospital. All patients with a confirmed or presumptive diagnosis of COVID-19 between March 1, 2020 and April 30, 2020 were considered potentially relevant. Infection with SARS-CoV-2 was confirmed by Roche Cobas 6800 polymerase chain reaction (PCR) in patients that were treated at the Mount Sinai Hospital. For patients admitted to other hospital systems, inclusion was based on external reporting and follow up testing confirmation. Similarly, outpatients that reported a positive COVID-19 test to our clinic (e.g. over the phone) were included in the analysis, awaiting collection of their formal test results. Anti-SARS-CoV-2 antibody testing was performed using an anti-IgG assay developed at Mount Sinai Health System Department of Pathology in collaboration with the Icahn School of Medicine at Mount Sinai Department of Microbiology under a Food and Drug Administration (FDA) Emergency Use Authorization. We reviewed clinical charts, nursing records, laboratory findings and radiological images for patients and obtained demographic data from the electronic medical records. Plasma levels of selected inflammatory cytokines, including IL-1β, IL-6, IL-8 and tumor necrosis factor-α (TNF-α), were assessed using the ELLA rapid detection enzyme-linked immunosorbent assay (ELISA) microfluidics platform and made available through the Mount Sinai data warehouse for hospitalized patients. Treatment response criteria were used as defined by the International Myeloma Working Group (IMWG)^20,21^. This retrospective study was approved by the institutional review board (IRB) of the Mount Sinai Hospital and is in compliance with the Declaration of Helsinki and International Conference on Harmonization Guidelines for Good Clinical Practice (IRB: GCO#: 11-1433).

### Statistical analysis

Continuous variables are presented as a median and interquartile range (IQR). Categorical variables are shown as percentage and absolute number of patients. Wherever two outcome groups are compared, Fisher’s exact test was used to determine significance and odds ratios (ORs) were reported for categorical variables and Mann-Whitney U test was used to determine significance for continuous variables. A two-sided alpha < 0.05 was considered statistically significant. All statistical analyses were done using R (version 3.6.1).

### Data availability

The datasets analyzed during the current study are not publicly available due to United States Federal Health Insurance Portability and Accountability Act (HIPAA) compliance, but a de-identified dataset may be available from the corresponding author on reasonable request.

## Results

### Baseline characteristics

Our cohort of 58 patients encompassed 52% males and had a median age of 67 years (IQR: 12.5 years), with 17% of patients older than 75 years (**Table 1**). The median body mass index (BMI) was 27.6 kg/m^2^ (with 37% of patients with a BMI > 30 kg/m^2^). The majority of patients reported being non-white (63%), with 13 (23%) patients of African American and 9 (16%) of Hispanic origin.

**Table 1:**
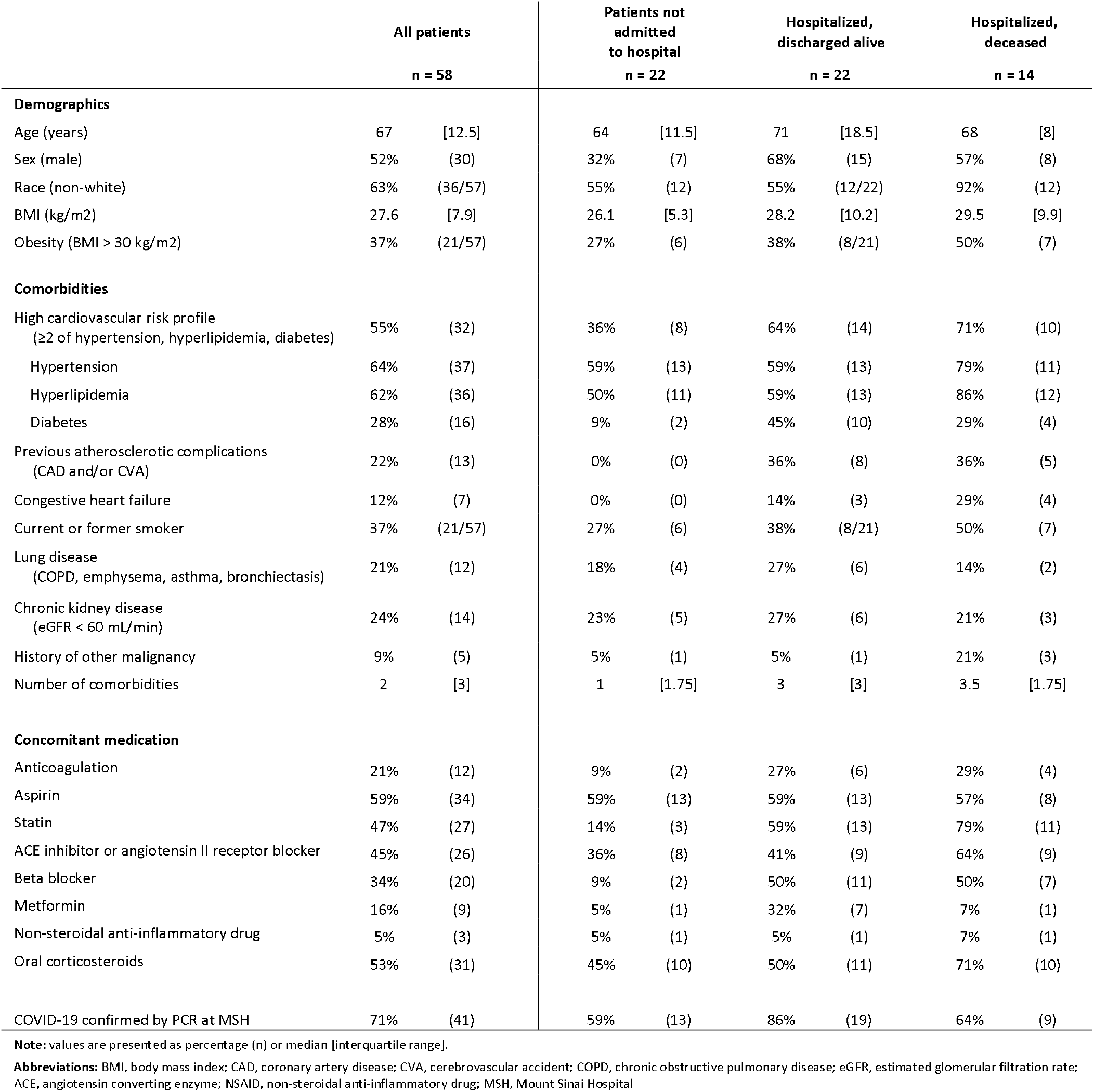
Demographics and baseline characteristics of patients.

The most common comorbidities were hypertension (64%), hyperlipidemia (62%), previous or active smoking (37%), diabetes mellitus type 2 (28%), chronic kidney disease (CKD, estimated glomerular filtration rate (eGFR) <60 mL/min) (24%) and lung disease (21%), including asthma or chronic obstructive lung disease (COPD). Thirty-two patients (55%) had a high-risk cardiovascular profile (defined as having ≥2 of the conditions: hypertension, hyperlipidemia and diabetes) and 13 (22%) had coronary artery disease (CAD) and/or cerebrovascular disease. Seven (12%) patients had congestive heart failure. Twelve (21%) patients were on therapeutic anticoagulation and 34 (59%) were on aspirin, while 26 (45%) patients were on an angiotensin-converting-enzyme (ACE) inhibitor or angiotensin II receptor blocker (ARB).

### Myeloma characteristics

The cohort included 54 MM and 4 SMM patients (**Table 2**). The median time from diagnosis to COVID-19 infection was 29.8 months (IQR: 44.2 months). MM patients had a median of 1.5 (IQR: 2) lines of therapy and 9 (17%) patients had more than 4 previous lines of treatment. Twenty-two (41%) patients had a prior autologous stem cell transplant (ASCT). The median age of patients with and without prior ASCT was 63.5 and 70 years, respectively. Of all patients, 27 (47%) had an Eastern Cooperative Oncology Group (ECOG) performance status of 0 at the time of COVID-19 infection. The most common myeloma subtype was IgG (59%) followed by IgA (19%), with light chain involvement in 33% of cases. High-risk cytogenetics were present in 22 (39%) patients, with 18 (33%) patients having an international staging system (ISS) of 1, while 14 (26%) and 10 (19%) patients had ISS 2 and 3, respectively, at time of MM diagnosis. At time of SARS-CoV-2 infection, 3 SMM and 8 MM patients were not on therapy. Among the remaining patients, 28 (48%) patients were being treated with daratumumab, 32 (55%) patients were on immunomodulatory drugs (IMiDs), 22 (38%) were on a proteasome inhibitor (PI), 5 (9%) were on venetoclax, and 30 (52%) patients were on concomitant corticosteroids.

**Table 2:** Myeloma disease characteristics of patients.

|  | All patients |  | Patients not<br>admitted<br>to hospital |  | Hospitalized,<br>discharged alive |  | Hospitalized,<br>deceased |  |
| --- | --- | --- | --- | --- | --- | --- | --- | --- |
|  | n = 58 |  | n = 22 |  | n = 22 |  | n = 14 |  |
| Disease characteristics |  |  |  |  |  |  |  |  |
| SMM | 7% | (4) | 9% | (2) | 5% | (1) | 7% | (1) |
| SMM/MM subtype |  |  |  |  |  |  |  |  |
| IgD | 2% | (1) | 5% | (1) | 0% | (0) | 0% | (0) |
| IgG | 59% | (34) | 59% | (13) | 64% | (14) | 50% | (7) |
| IgA | 19% | (11) | 23% | (5) | 14% | (3) | 21% | (3) |
| Light chain disease | 33% | (19) | 23% | (5) | 41% | (9) | 36% | (5) |
| Extramedullary disease history | 31% | (18) | 36% | (8) | 32% | (7) | 21% | (3) |
| ISS at diagnosis |  |  |  |  |  |  |  |  |
| 1 | 33% | (18/54) | 45% | (9/20) | 43% | (9/21) | 0% | (0/13) |
| 2 | 26% | (14/54) | 25% | (5/20) | 33% | (7/21) | 15% | (2/13) |
| 3 | 19% | (10/54) | 20% | (4/20) | 14% | (3/21) | 23% | (3/13) |
| not known | 22% | (12/54) | 10% | (2/20) | 10% | (2/21) | 62% | (8/13) |
| High risk cytogenetics | 39% | (22/56) | 33% | (7/21) | 41% | (9) | 46% | (6/13) |
| Time since MM diagnosis (months) | 29.8 | [44.2] | 44.8 | [38.7] | 27.2 | [55.8] | 28.6 | [23.6] |
| History of ASCT | 41% | (22/54) | 60% | (12/20) | 24% | (5/21) | 38% | (5/13) |
| Time since ASCT (days) | 985 | [1590] | 985 | [744] | 2148 | [2206] | 460 | [1734] |
| ASCT within last year | 9% | (5/54) | 15% | (3/20) | 0% | (0/21) | 15% | (2/13) |
| Lines of therapy (n) | 1.5 | [2] | 2 | [2] | 2 | [3] | 1 | [1] |
| More than 4 lines of treatment | 17% | (9/54) | 20% | (4/20) | 19% | (4/21) | 8% | (1/13) |
| ECOG 0 | 47% | (27) | 64% | (14) | 41% | (9) | 29% | (4) |
| Current response status* |  |  |  |  |  |  |  |  |
| sCR or CR | 26% | (15) | 45% | (10) | 14% | (3) | 14% | (2) |
| VGPR | 19% | (11) | 18% | (4) | 14% | (3) | 29% | (4) |
| PR | 22% | (13) | 18% | (4) | 23% | (5) | 29% | (4) |
| SD | 3% | (2) | 5% | (1) | 5% | (1) | 0% | (0) |
| PD | 16% | (9) | 5% | (1) | 23% | (5) | 21% | (3) |
| Not evaluable | 14% | (8) | 9% | (2) | 23% | (5) | 7% | (1) |
| Current MM treatment regimen |  |  |  |  |  |  |  |  |
| Contains CD38 mAb | 48% | (28) | 50% | (11) | 50% | (11) | 43% | (6) |
| Contains IMiD | 55% | (32) | 55% | (12) | 59% | (13) | 50% | (7) |
| Contains proteasome inhibitor | 38% | (22) | 27% | (6) | 41% | (9) | 50% | (7) |
| Contains corticosteroids | 52% | (30) | 45% | (10) | 50% | (11) | 64% | (9) |
| Contains venetoclax | 9% | (5) | 14% | (3) | 9% | (2) | 0% | (0) |
| No active treatment | 19% | (11) | 18% | (4) | 18% | (4) | 21% | (3) |
| Biochemical parameters<br>at last clinic visit before COVID-19 episode |  |  |  |  |  |  |  |  |
| Leukocyte count (x10e9/L) | 4.6 | [2.1] | 4.1 | [2.1] | 5.2 | [2.4] | 4.7 | [1.5] |
| Leukocytopenia (< 4 x 10e9/L) | 35% | (20/57) | 41% | (9) | 33% | (7/21) | 29% | (4) |
| Absolute neutrophil count (x10e9/L) | 2.6 | [2.2] | 2.4 | [1] | 3.4 | [3] | 2.4 | [1.4] |
| Neutropenia (< 2 x10e9/L) | 26% | (15/57) | 32% | (7) | 29% | (6/21) | 14% | (2) |
| Absolute lymphocyte count (x10e9/L) | 1 | [0.8] | 1.1 | [1.2] | 0.8 | [0.9] | 1.2 | [0.5] |
| Lymphocytopenia (< 0.5 x 10e9/L) | 12% | (7/57) | 0% | (0) | 33% | (7/21) | 0% | (0) |
| Serum free light chain ratio (involved/uninvolved) | 2.5 | [11.9] | 1.6 | [3.3] | 7.7 | [45.5] | 1.8 | [5.2] |
| M spike (g/dL) | 0 | [0.6] | 0 | [0.2] | 0.3 | [1.2] | 0.1 | [0.4] |
| IgG (mg/dL) | 805 | [736] | 1074.5 | [683.8] | 764 | [1121] | 727 | [496.8] |
| Hypogammaglobulinemia (IgG < 700 mg/dL) | 37% | (21/57) | 32% | (7) | 38% | (8/21) | 43% | (6) |
| Severe hypogammaglobulinemia (IgG < 400 mg/dL) | 11% | (6/57) | 0% | (0) | 10% | (2/21) | 29% | (4) |
| Immunoparesis | 89% | (51/57) | 82% | (18) | 95% | (20/21) | 93% | (13) |

**Note:** values are presented as percentage (n) or median [interquartile range].
**Abbreviations:** SMM, smoldering multiple myeloma; Ig, immunoglobulin; ISS, international staging system; MM, multiple myeloma; ASCT, autologous stem cell transplant; ECOG, Eastern Cooperative Oncology Group; sCR, stringent complete response; CR, complete response; VGPR, very good partial response; SD, stable disease; PD, progressive disease; mAb, monoclonal antibody; IMiD, immunomodulatory agent; \*according to IMWG criteria

The disease status at the time of SARS-CoV-2 infection included 15 (26%) patients in complete response (CR) or stringent CR (sCR), 11 (19%), 13 (22%) and 2 (3%) patients who had a very good partial response (VGPR), partial response (PR) and stable disease (SD), respectively, and 9 (16%) who had progressive disease (PD). Response status was not evaluable for 8 (14%) of patients (including 4 SMM patients and 1 newly diagnosed patient).

Biochemical parameters at the last routine clinic visit before presentation with COVID-19 were collected to determine if these steady-state parameters would provide insight into which patients are particularly vulnerable (**Table 2**). Twenty patients (35%) had leukopenia (<4 × 10^9^/L) and 7 (12%) lymphocytopenia (grade 3, <0.5 × 10^9^/L) at their last clinic visit. The monoclonal spike (M-spike) was undetectable in 31 (54%) patients. Median serum IgG level of all patients was 805 mg/dL (IQR: 736 mg/dL) and 37% (21/57) of patients had hypogammaglobulinemia (< 700 mg/dL), while 11% (6/57) of patients had severe (< 400 mg/dL) hypogammaglobulinemia. Immunoparesis, defined as a reduction in one or more of the uninvolved immunoglobulins below the lower limit of normal^22^, was present in 89% (51/57) of patients.

### Clinical course and biochemical parameters

The most common reported symptoms among all patients were fever (70%), cough (65%), and dyspnea (45%). Thirty-six patients were admitted at a hospital for inpatient care, 23 of which were admitted at our healthcare system and had both clinical and biochemical parameters available, as shown in **Table 3**. The median time between self-reported symptom onset and admission was 3 days. Among the 23 patients, 16 (70%) were febrile, and 11 (48%) were tachycardic with a heart rate >100 beats per minute (bpm) at time of presentation. Ten (43%) patients required immediate oxygen support: 7 needed a nasal cannula or non-rebreather mask, 1 needed high flow oxygen and 2 were immediately intubated and required mechanical ventilation.

**Table 3:** Clinical parameters, treatments and outcomes of patients hospitalized due to COVID-19 at Mount Sinai Hospital.

| *Subset of patients treated at Mount Sinai hospital for which full clinical data was available* | Hospitalized patients total |  | Hospitalized patients alive |  | Hospitalized patients deceased |  |
| --- | --- | --- | --- | --- | --- | --- |
|  | n = 23 |  | n = 16 |  | n = 7 |  |
| Clinical presentation |  |  |  |  |  |  |
| Fever | 70% | (16) | 69% | (11) | 71% | (5) |
| Systolic BP < 90 mmHg | 9% | (2) | 0% | (0) | 29% | (2) |
| MAP < 65 mmHg | 9% | (2) | 0% | (0) | 29% | (2) |
| Heart rate > 100 bpm | 48% | (11) | 56% | (9) | 29% | (2) |
| RR > 20/min | 26% | (6) | 19% | (3) | 43% | (3) |
| Oxygen requirement at presentation |  |  |  |  |  |  |
| None | 57% | (13) | 69% | (11) | 29% | (2) |
| Nasal cannula or NRB mask | 30% | (7) | 25% | (4) | 43% | (3) |
| High-flow oxygen, CPAP or BiPAP | 4% | (1) | 0% | (0) | 14% | (1) |
| Mechanical ventilation | 9% | (2) | 6% | (1) | 14% | (1) |
| Treatment initiated |  |  |  |  |  |  |
| Remdesivir | 4% | (1) | 6% | (1) | 0% | (0) |
| Hydroxychloroquine | 74% | (17) | 69% | (11) | 86% | (6) |
| Azithromycin | 74% | (17) | 75% | (12) | 71% | (5) |
| Other antibiotics | 83% | (19) | 81% | (13) | 86% | (6) |
| G-CSF | 26% | (6) | 38% | (6) | 0% | (0) |
| Therapeutic anticoagulation | 78% | (18) | 69% | (11) | 100% | (7) |
| Systemic corticosteroids | 43% | (10) | 31% | (5) | 71% | (5) |
| Anti-TNF | 4% | (1) | 0% | (0) | 14% | (1) |
| Anti-IL-1 | 9% | (2) | 0% | (0) | 29% | (2) |
| Anti-IL-6 | 17% | (4) | 13% | (2) | 29% | (2) |
| Selinexor | 22% | (5) | 25% | (4) | 14% | (1) |
| Convalescent plasma | 4% | (1) | 6% | (1) | 0% | (0) |
| Complications |  |  |  |  |  |  |
| Highest level of oxygen requirement |  |  |  |  |  |  |
| None | 26% | (6) | 31% | (5) | 14% | (1) |
| Nasal cannula or NRB mask | 39% | (9) | 56% | (9) | 0% | (0) |
| High-flow oxygen, CPAP or BiPAP | 13% | (3) | 6% | (1) | 29% | (2) |
| Mechanical ventilation | 22% | (5) | 6% | (1) | 57% | (4) |
| ICU | 30% | (7) | 6% | (1) | 86% | (6) |
| Acute kidney injury | 52% | (12) | 38% | (6) | 86% | (6) |
| Shock | 30% | (7) | 0% | (0) | 100% | (7) |
| Sepsis or HAP/VAP | 9% | (2) | 6% | (1) | 14% | (1) |
| C. difficile infection | 4% | (1) | 6% | (1) | 0% | (0) |
| Cardiac complication | 30% | (7) | 19% | (3) | 57% | (4) |
| Worst biochemical parameters |  |  |  |  |  |  |
| CRP | 151.5 | [178.1] | 144.9 | [107.7] | 294.1 | [132.9] |
| Total leukocyte count (x10e9/L) (lowest) | 3.3 | [2.6] | 2.9 | [2.4] | 4.3 | [8.4] |
| Absolute lymphocyte count (x10e9/L) (lowest) | 0.3 | [0.4] | 0.4 | [0.5] | 0.2 | [0.2] |
| Creatinine (mg/dL) | 1.6 | [2.4] | 1.1 | [1.4] | 2.7 | [2] |
| Procalcitonin (ng/mL) | 0.8 | [2.1] | 0.5 | [0.8] | 2.7 | [9.9] |
| Ferritin (µg/L) | 2537 | [2578] | 1282 | [2170.5] | 3409 | [2018] |
| Fibrinogen (mg/dL) | 667 | [167] | 646 | [209.5] | 673.5 | [43.8] |
| D-dimer (mg/L) | 2.5 | [14.3] | 2 | [2.6] | 20 | [7.6] |
| LDH (U/L) | 531.5 | [515.8] | 478 | [329] | 739 | [344] |
| ALT (U/L) | 61 | [58.5] | 43.5 | [52.8] | 79 | [32] |
| AST (U/L) | 78 | [62.5] | 49.5 | [54.3] | 101 | [51.5] |
| IL-1β (pg/mL) | 0.5 | [0.9] | 0.5 | [0.8] | 0.5 | [0.9] |
| IL-6 (pg/mL) | 128.5 | [211.6] | 119.3 | [102.4] | 296.8 | [821.2] |
| IL-8 (pg/mL) | 65.4 | [62.3] | 46.6 | [67.6] | 137 | [89.6] |
| TNF-alfa (pg/mL) | 28.7 | [11] | 29.4 | [8.4] | 21.3 | [15.2] |

**Note:** values are presented as percentage (n) or median [interquartile range].
**Abbreviations:** BP, blood pressure; MAP, mean arterial pressure; SpO<sub>2</sub>, oxygen saturation; RR, respiratory rate; NRB, non-rebreather; CPAP, continuous positive airway pressure; BiPAP, bi-level positive airway pressure; G-CSF, granulocyte colony stimulating factor; TNF, tumor necrosis factor; IL, interleukin; ICU, intensive care unit; HAP, hospital-acquired pneumonia; VAP, ventilator-associated pneumonia; CRP, C-reactive protein; LDH, lactate dehydrogenase; ALT, alanine aminotransferase; AST, aspartate aminotransferase

During their hospital stay, 22 (95%) patients developed fever, 18 (78%) tachycardia (>100 bpm) and 18 (78%) hypoxemia (SpO2 < 93%). Five (22%) patients never required supplemental oxygen and 10 (40%) needed a nasal cannula or non-rebreather mask at some point during hospitalization. Four (17%) patients were treated with high-flow oxygen, continuous positive airway pressure (CPAP) or bi-level positive airway pressure (BiPAP) machines and five (22%) were eventually intubated. Seven (30%) patients required intensive care unit (ICU) care during their hospitalization. The median length of stay was 22 days. Of the 23 patients admitted to our hospital with COVID-19, seven (30%) died. When we consider the total hospitalized cohort (36 patients, i.e. including patients admitted at other hospitals), the mortality rate was 39% (14 patients). All 14 deaths were due to COVID-19. There were no deaths reported in patients who were not hospitalized among the total cohort.

Patients presented with multiple elevated inflammatory markers, including CRP (median: 89 mg/L), ferritin (median: 595 µg/L), IL-6 (median: 82.4 pg/mL), whereas procalcitonin was normal (median: 0.2 ng/mL). Leukocytes were not elevated (median: 4.3×10^9^/L) and the absolute lymphocyte count was low (median 0.6 × 10^9^/L) whereas absolute neutrophil count was within normal range (median 3.6 × 10^9^/L). On initial presentation, lactate dehydrogenase (LDH, median 249.5 U/L), fibrinogen (median 600 mg/dL) and D-dimer (median 1.2 mg/L) were elevated but transaminases were normal (median aspartate aminotransferase (AST) and alanine aminotransferase (ALT): 24 U/L and 20 U/L, respectively). Peak levels for these markers are shown in **Table 3** and temporal trends for a subset of patients are illustrated in **Supplementary Figure S1**. CRP and ferritin peaked early (within the first 10 days of hospitalization) and subsequently demonstrated a downward trend over time, with a slower decline in ferritin levels notable in patients that eventually died. D-dimer level was transiently elevated in some patients but was persistently and progressively elevated in all patients that eventually died. The inflammatory cytokines IL-1β, IL-6, IL-8 and TNF-α, were assessed over the duration of hospitalization at the discretion of the treating physician. Peak levels for IL-6, IL-8 and TNF-α were elevated (median: 128.5 pg/mL, 65.4 pg/mL and 28.7 pg/mL, respectively) and levels for IL-1β were generally low (median: 0.5pg/mL).

We describe COVID-19 management comprehensively for the 23 patients hospitalized at our center and their outcome (**Table 3**). One (4%) patient received remdesivir, 17 (74%) patients received hydroxychloroquine and 17 (74%) patients received azithromycin. Nineteen (82%) patients were treated with other antibiotics, most commonly a beta-lactam antibiotic ± vancomycin (n = 16) for presumed bacterial superinfection. Six (26%) patients received granulocyte colony stimulating factor (G-CSF) and 18 (78%) patients received therapeutic anticoagulation, 13 of which had not been on full anticoagulation before COVID-19. Patients were treated with a direct oral anticoagulant (DOAC, n = 3), therapeutic doses of enoxaparin (n = 8) or a heparin drip (n = 2). There were no major bleeding events. Ten (43%) patients were given systemic corticosteroids. One (4%) patient was treated with convalescent plasma. Anti-TNFα, anti-IL-6 and anti-IL-1 therapy was initiated in 1 (4%), 4 (17%) and 2 (9%) patients, respectively. Five patients (22%) were given low dose selinexor, a selective inhibitor of nuclear export, for its presumed activity against virus host protein interaction^23^ and to counter amplification of pro-inflammatory signaling^24^.

### Antibody serology and repeat PCR testing

We collected data on antibody testing and follow up PCR testing for patients. As of May 28, 2020, 96% (22/23) of patients have developed antibodies against SARS-CoV-2 at a median time of 32 (range 6-50) days since COVID-19 diagnosis. Titers ranged from 1:160 to 1:2880, with 74% (17/23) exhibiting the most significant titer level of 1:2880, 9% (2/23) with 1:960, 9% (2/23) with 1:320, and 4% (1/23) with 1:160. Antibody titer did not correlate with COVID-19 severity, yet we observed that all 5 MM patients with low titers (< 1:2880) had hypogammaglobulinemia. The one patient who did not develop any antibodies had SMM and was tested 27 days after initial diagnosis. So far, 27 patients have undergone repeat PCR testing; 74% (20/27) are negative and median time to PCR negativity was 43 (range 19-68) days from initial positive PCR. Among the 22 patients with positive antibody titers, 19 patients had repeat PCR swab and 3 remained positive while 16 were negative.

### Clinical associations

In a univariate analysis on all patients, we found that the following variables were significantly associated with hospitalization, as shown in **Table 4**: age over 70 (OR 7.74, p = 0.007), male sex (OR 3.70, p = 0.030), diabetes mellitus type 2 (OR 6.18, p = 0.016), high cardiovascular risk profile (OR 3.42, p = 0.032), history of CAD (OR ∞, p = 0.009), history of CHF (OR ∞, p = 0.037), use of statins (OR = 12.10, p < 0.001) and use of beta blockers (OR 9.63, p = 0.002). We also noted significant associations between hospitalization status and grade 3 lymphocytopenia (OR ∞, p = 0.036) at the last clinic visit prior to COVID-19 infection. Patients who had not achieved complete remission or stringent complete remission were at increased risk of hospitalization (p = 0.013).

**Table 4:**
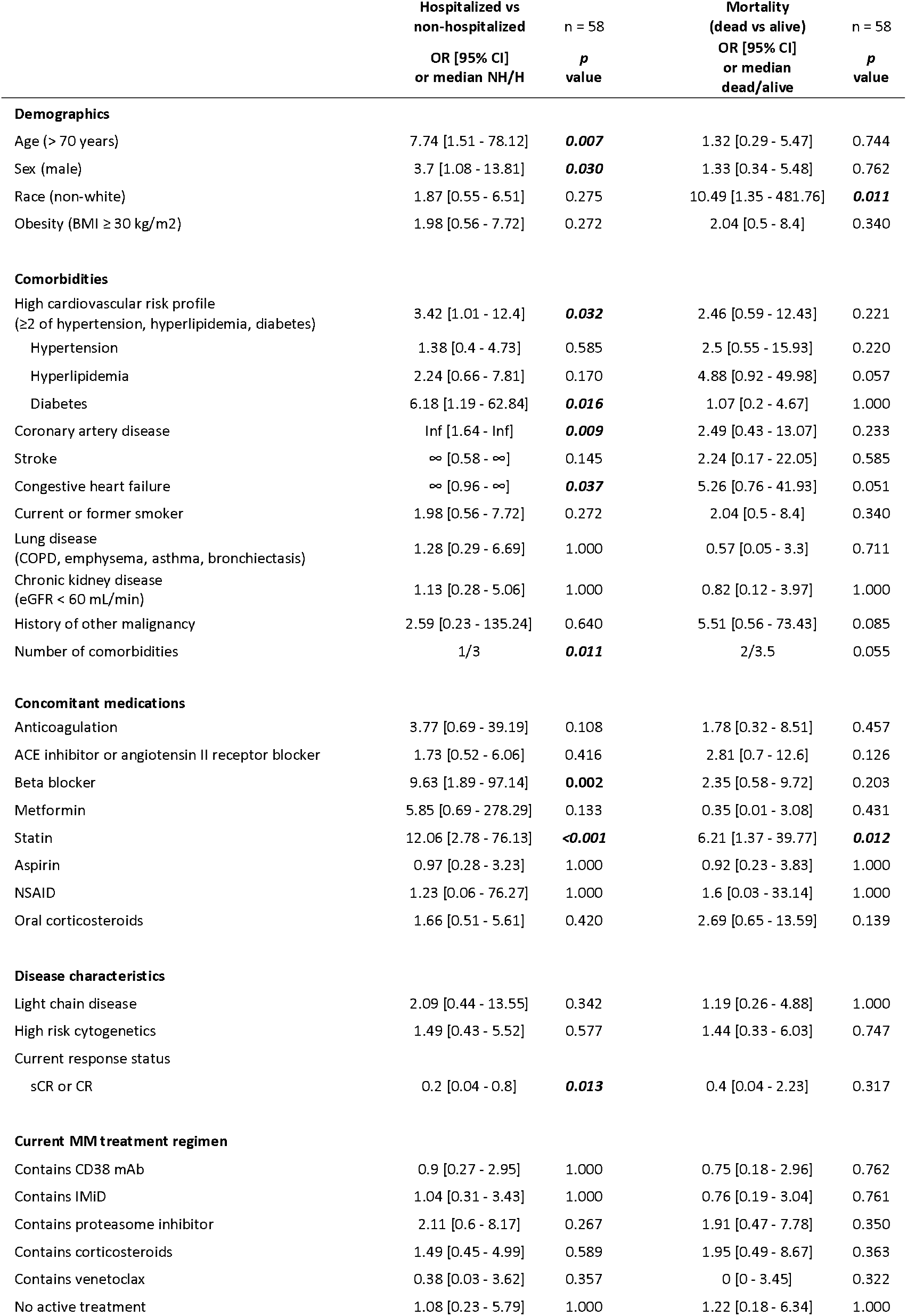
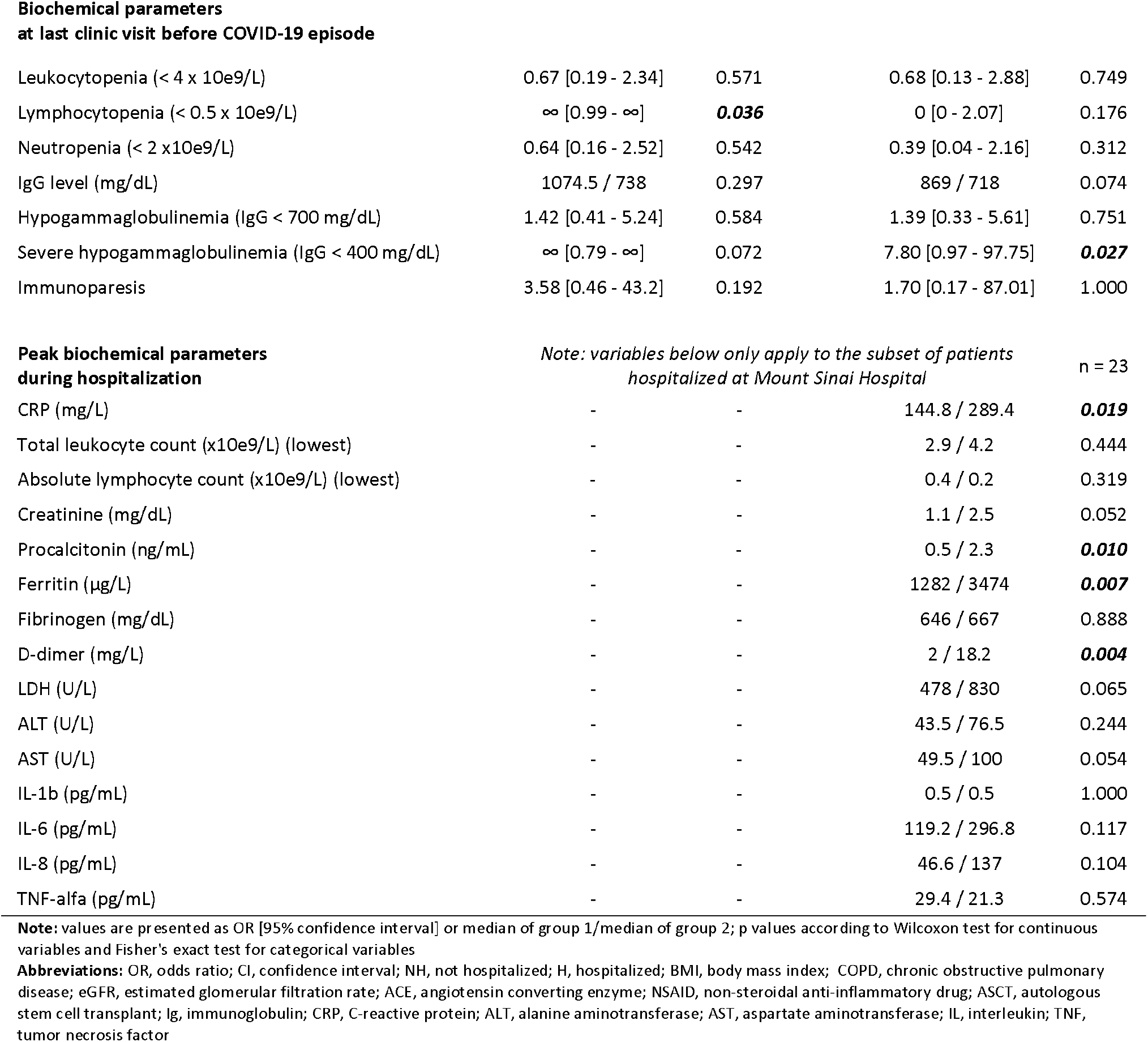
Univariate associations of selected variables with risk of hospitalization and mortality.

|  | Hospitalized vs<br>non-hospitalized<br>OR [95% CI]<br>or median NH/H | n = 58<br>*p*<br>value | Mortality<br>(dead vs alive)<br>OR [95% CI]<br>or median<br>dead/alive | n = 58<br>*p*<br>value |
| --- | --- | --- | --- | --- |
| **Demographics** |  |  |  |  |
| Age (> 70 years) | 7.74 [1.51 - 78.12] | **0.007** | 1.32 [0.29 - 5.47] | 0.744 |
| Sex (male) | 3.7 [1.08 - 13.81] | **0.030** | 1.33 [0.34 - 5.48] | 0.762 |
| Race (non-white) | 1.87 [0.55 - 6.51] | 0.275 | 10.49 [1.35 - 481.76] | **0.011** |
| Obesity (BMI ≥ 30 kg/m<sup>2</sup>) | 1.98 [0.56 - 7.72] | 0.272 | 2.04 [0.5 - 8.4] | 0.340 |
| **Comorbidities** |  |  |  |  |
| High cardiovascular risk profile<br>(≥2 of hypertension, hyperlipidemia, diabetes) | 3.42 [1.01 - 12.4] | **0.032** | 2.46 [0.59 - 12.43] | 0.221 |
| Hypertension | 1.38 [0.4 - 4.73] | 0.585 | 2.5 [0.55 - 15.93] | 0.220 |
| Hyperlipidemia | 2.24 [0.66 - 7.81] | 0.170 | 4.88 [0.92 - 49.98] | 0.057 |
| Diabetes | 6.18 [1.19 - 62.84] | **0.016** | 1.07 [0.2 - 4.67] | 1.000 |
| Coronary artery disease | Inf [1.64 - Inf] | **0.009** | 2.49 [0.43 - 13.07] | 0.233 |
| Stroke | ∞ [0.58 - ∞] | 0.145 | 2.24 [0.17 - 22.05] | 0.585 |
| Congestive heart failure | ∞ [0.96 - ∞] | **0.037** | 5.26 [0.76 - 41.93] | 0.051 |
| Current or former smoker | 1.98 [0.56 - 7.72] | 0.272 | 2.04 [0.5 - 8.4] | 0.340 |
| Lung disease<br>(COPD, emphysema, asthma, bronchiectasis) | 1.28 [0.29 - 6.69] | 1.000 | 0.57 [0.05 - 3.3] | 0.711 |
| Chronic kidney disease<br>(eGFR < 60 mL/min) | 1.13 [0.28 - 5.06] | 1.000 | 0.82 [0.12 - 3.97] | 1.000 |
| History of other malignancy | 2.59 [0.23 - 135.24] | 0.640 | 5.51 [0.56 - 73.43] | 0.085 |
| Number of comorbidities | 1/3 | **0.011** | 2/3.5 | 0.055 |
| **Concomitant medications** |  |  |  |  |
| Anticoagulation | 3.77 [0.69 - 39.19] | 0.108 | 1.78 [0.32 - 8.51] | 0.457 |
| ACE inhibitor or angiotensin II receptor blocker | 1.73 [0.52 - 6.06] | 0.416 | 2.81 [0.7 - 12.6] | 0.126 |
| Beta blocker | 9.63 [1.89 - 97.14] | **0.002** | 2.35 [0.58 - 9.72] | 0.203 |
| Metformin | 5.85 [0.69 - 278.29] | 0.133 | 0.35 [0.01 - 3.08] | 0.431 |
| Statin | 12.06 [2.78 - 76.13] | **<0.001** | 6.21 [1.37 - 39.77] | **0.012** |
| Aspirin | 0.97 [0.28 - 3.23] | 1.000 | 0.92 [0.23 - 3.83] | 1.000 |
| NSAID | 1.23 [0.06 - 76.27] | 1.000 | 1.6 [0.03 - 33.14] | 1.000 |
| Oral corticosteroids | 1.66 [0.51 - 5.61] | 0.420 | 2.69 [0.65 - 13.59] | 0.139 |
| **Disease characteristics** |  |  |  |  |
| Light chain disease | 2.09 [0.44 - 13.55] | 0.342 | 1.19 [0.26 - 4.88] | 1.000 |
| High risk cytogenetics | 1.49 [0.43 - 5.52] | 0.577 | 1.44 [0.33 - 6.03] | 0.747 |
| Current response status<br>sCR or CR | 0.2 [0.04 - 0.8] | **0.013** | 0.4 [0.04 - 2.23] | 0.317 |
| **Current MM treatment regimen** |  |  |  |  |
| Contains CD38 mAb | 0.9 [0.27 - 2.95] | 1.000 | 0.75 [0.18 - 2.96] | 0.762 |
| Contains IMiD | 1.04 [0.31 - 3.43] | 1.000 | 0.76 [0.19 - 3.04] | 0.761 |
| Contains proteasome inhibitor | 2.11 [0.6 - 8.17] | 0.267 | 1.91 [0.47 - 7.78] | 0.350 |
| Contains corticosteroids | 1.49 [0.45 - 4.99] | 0.589 | 1.95 [0.49 - 8.67] | 0.363 |
| Contains venetoclax | 0.38 [0.03 - 3.62] | 0.357 | 0 [0 - 3.45] | 0.322 |
| No active treatment | 1.08 [0.23 - 5.79] | 1.000 | 1.22 [0.18 - 6.34] | 1.000 |
| **Biochemical parameters at last clinic visit before COVID-19 episode** |  |  |  |  |
| Leukocytopenia (< 4 x 10e9/L) | 0.67 [0.19 - 2.34] | 0.571 | 0.68 [0.13 - 2.88] | 0.749 |
| Lymphocytopenia (< 0.5 x 10e9/L) | ∞ [0.99 - ∞] | **0.036** | 0 [0 - 2.07] | 0.176 |
| Neutropenia (< 2 x10e9/L) | 0.64 [0.16 - 2.52] | 0.542 | 0.39 [0.04 - 2.16] | 0.312 |
| IgG level (mg/dL) | 1074.5 / 738 | 0.297 | 869 / 718 | 0.074 |
| Hypogammaglobulinemia (IgG < 700 mg/dL) | 1.42 [0.41 - 5.24] | 0.584 | 1.39 [0.33 - 5.61] | 0.751 |
| Severe hypogammaglobulinemia (IgG < 400 mg/dL) | ∞ [0.79 - ∞] | 0.072 | 7.80 [0.97 - 97.75] | **0.027** |
| Immunoparesis | 3.58 [0.46 - 43.2] | 0.192 | 1.70 [0.17 - 87.01] | 1.000 |
| CRP (mg/L) | - | - | 144.8 / 289.4 | **0.019** |
| Total leukocyte count (x10e9/L) (lowest) | - | - | 2.9 / 4.2 | 0.444 |
| Absolute lymphocyte count (x10e9/L) (lowest) | - | - | 0.4 / 0.2 | 0.319 |
| Creatinine (mg/dL) | - | - | 1.1 / 2.5 | 0.052 |
| Procalcitonin (ng/mL) | - | - | 0.5 / 2.3 | **0.010** |
| Ferritin (µg/L) | - | - | 1282 / 3474 | **0.007** |
| Fibrinogen (mg/dL) | - | - | 646 / 667 | 0.888 |
| D-dimer (mg/L) | - | - | 2 / 18.2 | **0.004** |
| LDH (U/L) | - | - | 478 / 830 | 0.065 |
| ALT (U/L) | - | - | 43.5 / 76.5 | 0.244 |
| AST (U/L) | - | - | 49.5 / 100 | 0.054 |
| IL-1b (pg/mL) | - | - | 0.5 / 0.5 | 1.000 |
| IL-6 (pg/mL) | - | - | 119.2 / 296.8 | 0.117 |
| IL-8 (pg/mL) | - | - | 46.6 / 137 | 0.104 |
| TNF-alfa (pg/mL) | - | - | 29.4 / 21.3 | 0.574 |

**Note:** values are presented as OR [95% confidence interval] or median of group 1/median of group 2; p values according to Wilcoxon test for continuous variables and Fisher's exact test for categorical variables
**Abbreviations:** OR, odds ratio; CI, confidence interval; NH, not hospitalized; H, hospitalized; BMI, body mass index; COPD, chronic obstructive pulmonary disease; eGFR, estimated glomerular filtration rate; ACE, angiotensin converting enzyme; NSAID, non-steroidal anti-inflammatory drug; ASCT, autologous stem cell transplant; Ig, immunoglobulin; CRP, C-reactive protein; ALT, alanine aminotransferase; AST, aspartate aminotransferase; IL, interleukin; TNF, tumor necrosis factor

Similarly, for hospitalized patients, using a univariate approach, we found statistically significant association between mortality and these variables: non-white race (OR 10.49, p = 0.011), statin use (OR 6.21, p = 0. 012), severe hypogammaglobulinemia (OR 7.80, p = 0.027), and higher peak levels of D-dimer (p = 0.004), ferritin (p = 0.007), procalcitonin (p = 0.010), and CRP (p = 0.019). The full list of associations is shown in **Table 4**.

## Discussion

Situated in the heart of New York City, our cancer center at Mount Sinai Hospital bore witness to the immense disruption of healthcare caused by COVID-19. During the initial phase of the pandemic, the goal was to keep patients at home following federal and state guidelines of isolation, social distancing, and strict hand hygiene^25-27^. Patients were switched to all oral regimens if possible or had delayed therapy depending on perceived risk of need for therapy to control myeloma versus exposure to SARS-CoV-2. Yet community transmission of SARS-CoV-2 during the pandemic was inevitable.

There were no deaths among myeloma patients with milder symptoms who were managed entirely as outpatients with COVID-19 in this cohort. The mortality rates of the overall cohort (n = 58), MM patients admitted to Mount Sinai Hospital (n = 23), and all admitted MM patients (n = 36) were 24%, 30%, and 39% respectively. These figures are in line with the overall mortality seen in New York, where the estimated mortality among hospitalized patients over 45 years old is 37% as of May 25, 2020^1,28^. Interestingly the mortality among our cohort of MM patients was lower than the 54.6% seen in a mixed cohort of MM patients treated in Britain^29^. We acknowledge that the apparent mortality differences between different countries and health systems may be affected by the local epidemiology, hospitalization and resource utilization rates and potential differences in escalation of care. However, in both of these populations, there appeared to be a trend toward increased mortality in patients of non-white/Caucasian background. This has been consistently seen in the United States, where death rates for COVID-19 are several fold higher in patients of Black and Hispanic origins^30-32^.

MM specific disease characteristics and the type of MM treatment were not associated with increased mortality. In contrast, we observed that age and cardiovascular risk factors (diabetes, CAD, CHF) were significantly associated with patient hospitalization for COVID-19. The data from our cohort showed that non-white background, severe (< 400 mg/dL) hypogammaglobulinemia, and statin use were significantly associated with mortality. This information would indicate that during the post pandemic phase, we do not have to change the management of myeloma patients. However, earlier diagnosis of COVID-19 and prompt intervention especially for the vulnerable population identified above is warranted to reduce the risk of mortality. As we reopen and move forward into a post-COVID-19 era, we will need to remain vigilant, particularly for select patient groups, and await effective COVID-19 treatments while balancing the need to manage patients’ myeloma.

We were able to capture the evolution of inflammatory markers for patients who were admitted to the inpatient service and we found a significant association with mortality in patients who had elevated D-dimer, CRP, or ferritin. Many COVID-19 patients treated at our institution also received a rapid panel for cytokine testing as part of a larger study to characterize the inflammatory profile of COVID-19 illness. Among our cohort of hospitalized patients, those who died appeared to exhibit rather elevated pro-inflammatory cytokines, consistent in principle with what was seen in a large COVID-19 cohort analyzed at the Mount Sinai Health System^33^. It is possible that a CRS-like syndrome, similar though not identical to one seen in MM patients treated with CAR-T^14,34^ and bispecific antibodies^35,36^, occurs in a significant portion of MM patients afflicted with COVID-19. Various agents including but not limited to anti-IL-6^39^ monoclonal antibodies and JAK inhibitors^40^ are presently under investigation to address potential components of immune dysregulation in COVID-19. We noted that patients who died from COVID-19 had alarmingly elevated D-dimer levels compared to survivors (median of 18.24 mg/L vs 1.96 mg/L). Emerging research suggests that the overwhelming immune activation during SARS-CoV-2 infection is a potent catalyst for significant arterial and venous thromboembolism leading to strokes and pulmonary emboli^16,37^, and serum pro-inflammatory cytokines including IL-1β, TNF-α, and IL-6 have been tied to endothelial damage underlying thrombus formation seen in COVID-19^38^. To counter this possibility, a large majority of patients admitted to our institution in this cohort received therapeutic anticoagulation and none suffered bleeding events. Our data regarding inflammatory markers raises the question if the process driving severe D-dimer elevation in MM patients with COVID-19 is the same or is separate from the CRS-like process seen in many COVID-19 patients.

Data on the persistence of SARS-CoV-2 by PCR and development of specific antibody response to the virus in potentially immunocompromised cancer patients have thus far been lacking. A significant majority of tested patients among this cohort cleared infection by PCR and developed antibodies despite a very high proportion of patients who fit the definition of classical myeloma-associated immunoparesis. Immunoparesis alone was not significantly associated with hospitalization or mortality and importantly did not appear to affect the development of anti-SARS-CoV-2 antibodies. Looking forward, we will need to determine if development of antibodies confers protection against reinfection.

This study has the limitations of single institution, retrospective reporting of a smaller cohort of patients. Serological data were not available for a minority of the patients who were hospitalized at outside institutions. The observations reported here have to be confirmed by a larger series of data collected from multiple institutions and such efforts are underway. Few patients received COVID-19 directed treatment on clinical trials. The role of recently emergency approved anti-viral agent remdesivir or convalescent plasma should be explored in the high-risk population with myeloma.

## Conclusions

In this study of patients treated for myeloma at the Mount Sinai Hospital, we provide a detailed analysis of a cohort of 58 MM and SMM patients who developed COVID-19. Although several demographic factors and comorbidities increased risk of hospitalization and mortality, myeloma therapy and immunoparesis did not influence outcomes. In fact, survival was comparable to the overall population of New York during the pandemic, and patients generally mounted a significant antibody response to SARS-CoV-2. The data herein supports the need to maintain proactive management of MM patients by balancing their need for therapy with the increased risk of hospitalization and death in a subset of MM patients with COVID-19.

## Abbreviations

ACE: angiotensin-converting-enzyme
ALT: alanine aminotransferase
ARB: angiotensin II receptor blocker
ASCT: autologous stem cell transplant
AST: aspartate aminotransferase
BiPAP: bi-level positive airway pressure
BMI: body mass index
bpm: beats per minute
CAD: coronary artery disease
CAR: chimeric antigen receptor
CHF: congestive heart failure
CKD: chronic kidney disease
COPD: chronic obstructive lung disease
COVID-19: coronavirus disease 2019
CPAP: continuous positive airway pressure
CR: complete response
CRS: cytokine release syndrome
CRP: C-reactive protein
DOAC: direct oral anticoagulant
ECOG: Eastern Cooperative Oncology Group
eGFR: estimated glomerular filtration rate
ELISA: enzyme-linked immunosorbent assay
G-CSF: granulocyte colony stimulating factor
HIPAA: Health Insurance Portability and Accountability Act
IMWG: International Myeloma Working Group
ICU: intensive care unit
IL: interleukin
IMiD: immunomodulatory drug
IRB: institutional review board
IQR: interquartile range
ISS: international staging system
LDH: lactate dehydrogenase
MM: Multiple myeloma
OR: odd’s ratio
PCR: polymerase chain reaction
PD: progressive disease
PI: proteasome inhibitor
PR: partial response
SARS-CoV-2: Severe Acute Respiratory Syndrome Coronavirus 2
sCR: stringent complete response
SD: stable disease
SMM: smoldering multiple myeloma
TNF-α: tumor necrosis factor-α
VGPR: very good partial response.

## Author Contributions

Conceptualization, BW, OVO, THM, SJ, SP, and DM; Methodology, OVO; Investigation, BW, OVO, THM, DV and SG; Writing – Original Draft, BW, OVO, THM; Writing – Review & Editing, all authors; Funding Acquisition, SJ.; Resources, MM, SJ, SP and DM; Supervision, MM, SJ, SP and DM.

## Acknowledgements

We would like to acknowledge all the staff and families for the selfless efforts in caring for patients who developed COVID-19, and the strength and courage of all patients affected by the pandemic.

## Supplementary Figures

**Figure S1:**
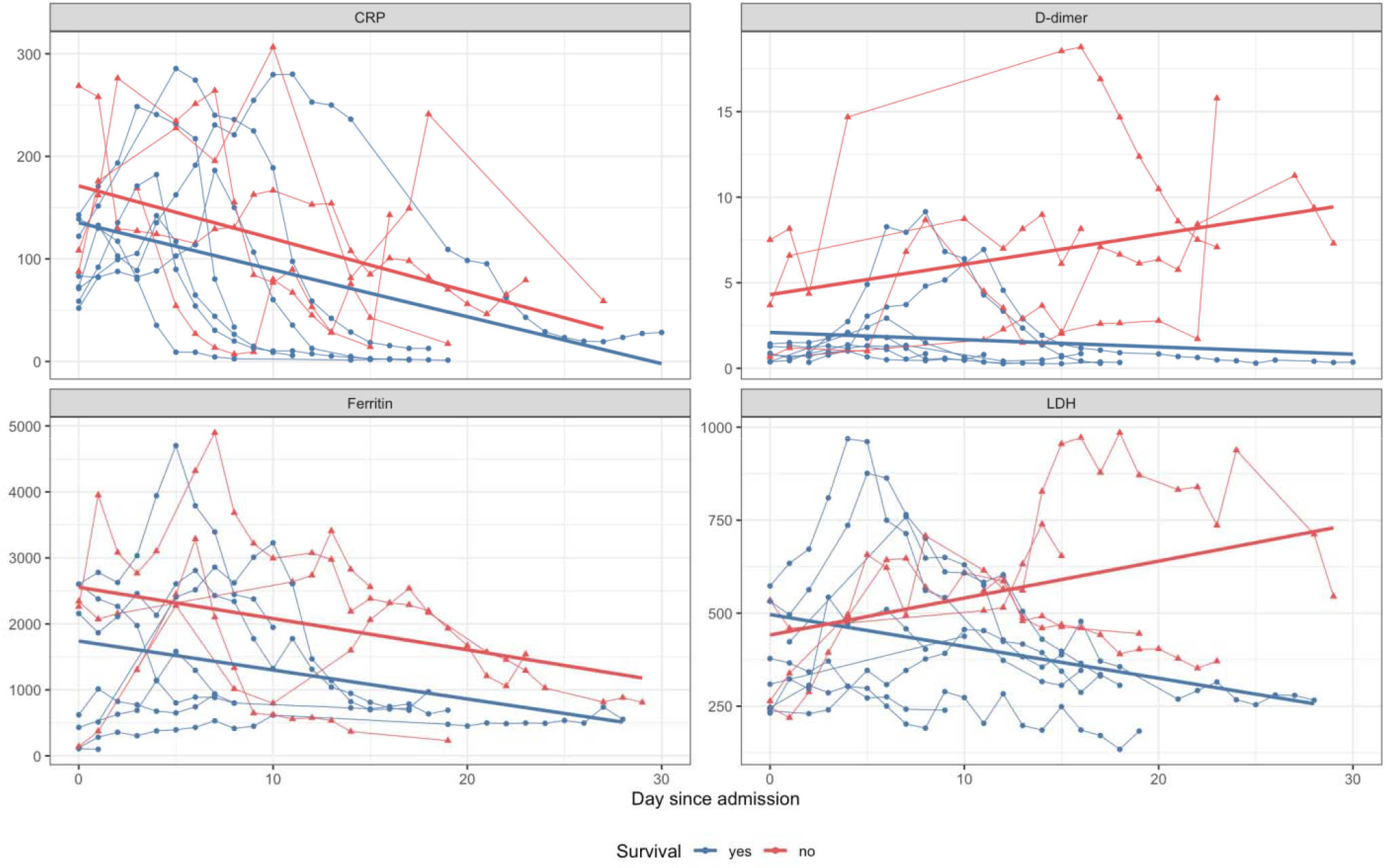
Evolution of selected inflammatory biomarkers in a subset of patients (n = 12) hospitalized at the Mount Sinai Hospital for which the data was available. Different measurements from the same patient are connected. A linear regression line is plotted for the subgroup of patients that survived (blue, n = 8) and died (red, n = 4), respectively.

